# Experiences with Primary Care in the State of Utah for Gender Diverse People: Surveying the Difficulties in Accessing Healthcare

**DOI:** 10.1101/2020.12.28.20248879

**Authors:** Bita Tristani-Firouzi, Joanne Rolls, Nicole L. Mihalopoulos, Cori A. Agarwal

## Abstract

**Background:** Transgender and non-binary communities continue to be underserved in healthcare. This study seeks to better understand the barriers and difficulties faced by transgender and non-binary patients in accessing primary care and hormone therapy in Utah.

**Methods:** An online survey was developed for transgender and non-binary identifying adults and was advertised via social media and the University of Utah Hospital’s website.

**Results:** There were 123 respondents from Utah including 39 trans women, 49 trans men, and 35 non-binary individuals. The age ranged from 18-67 (average 30 years), and 93% were Caucasian. The majority (84%) were insured, yet 67% of respondents reported difficulty accessing primary care. Fear of discrimination and being unable to find trans-friendly providers were reported as the two largest barriers. Non-binary respondents reported fear of discrimination as a barrier to primary care at the highest percentage (92%). Nearly 3 in 4 respondents who have hormone therapy reported difficulty paying for it. One in four trans women reported accessing hormones online or from a friend.

**Conclusion:** Utah is currently drastically underequipped to provide for the healthcare needs of transgender and non-binary communities. There needs to be an increase in trans-friendly primary care providers to curb discrimination. More resources and efforts must go into training primary care providers with necessary knowledge to properly serve transgender and non-binary patients. Finally, clear anti-discrimination laws are needed for insurance companies to reduce the financial barrier to transgender health services in Utah.

## Background

Transgender and non-binary individuals face considerable challenges when attempting to access healthcare. Discrimination in the healthcare sector can take the form of not using a patient’s chosen name or pronouns, to refusal of care completely.^1^ This treatment and stigma have led many transgender and non-binary individuals to avoid seeking healthcare altogether.^2,3^ Discrimination and stigma from healthcare is a contributing factor to health discrepancies between the trans population compared to the cis population.^4^ Transition-related healthcare, such as hormone therapy, is often accessed through primary care providers. Systematic review of studies has indicated that hormone therapy improves quality of life and psychological functioning for transgender and non-binary people.^5,6^ Cost is a major barrier to hormone therapy. While some insurance companies cover these costs, they are not required to do so, and often create barriers to coverage.^7^ Furthermore, lack of access to trans-friendly and trans-competent providers is a major barrier to transgender and non-binary nationally.^8^ While numerous studies have looked into the barriers to primary care and transition related care, there is a paucity of data specifically focusing on the diverse array of gender identities. Several studies have reported on experiences with the healthcare system for trans people in liberal cities and states,^9,10^ but data from conservative regions is lacking. Utah is a unique state, in that it is home to a significant percentage of transgender and non-binary individuals despite it being one of the most conservative states in the U.S.^11^ This paper focuses on the experiences of transgender and non-binary people with accessing primary care in the state of Utah across a variety of gender identities.

## Methods

An anonymous, nationally promoted survey of transgender and non-binary identifying individuals was developed and performed using a secure, web-based survey tool (REDCap) between January 2018 to October 2018 and was created specifically for this study (Supplemental File 1). The IRB approved the study. Inclusion criteria included (1) identifying as transgender or non-binary; (2) being 18 years of age or older; (3) residing in the United States; and (4) completing the survey. Informed consent was asked at the beginning of the survey. The survey included 146 multiple choice and free response questions surrounding demographics, insurance, employment, education, mental health, and questions regarding access and experiences with primary care. The survey was advertised on social media, the University of Utah hospital website, and through Planned Parenthood of Utah. Participants filled out the survey on their own time without a facilitator or spectator. Statistical averages were taken from multiple choice questions via averaging. Only participants who selected state of residence in Utah were analyzed. Participants were asked for their sex assigned at birth and then asked to select their gender identity from a list including trans woman, trans man, non-binary, genderqueer, agender, transfeminine, transmasculine, and an open response named “other.” For the purposes of this paper, we created three groups for sub-analysis: 1- trans men, 2- trans women, and a third group, non-binary, in which we combined all gender identity groups that did not identify as one of the binary options. While it would have been preferable to evaluate each gender identity independently, the small sample sizes did not allow for meaningful sub-analysis of each group.

## Results

A total of 1,092 responses from across the United States were collected. After eliminating partial responses or duplicates, a total of 887 individuals were included in the study who represented 49 states. There were 123 respondents from Utah, including 39 trans women, 49 trans men, and 35 non-binary individuals. From the Utah data, the average age was 30 years with a range of 18 to 67 years. The vast majority of respondents identified as Caucasian (93%). The majority of the participants either had some college education (including students) or a college degree and beyond (80%). Non-binary respondents and trans women reported the highest level of education (49% having a college degree or higher) (Table 1). Approximately half of the respondents indicated they had full time employment. On average, 12% selected they were unemployed, with non-binary participants having the lowest percentage of unemployment (7%). Most of the participants had health insurance (84%). Trans women, on average, had the lowest percentage of insurance (67%) whereas nearly all the non-binary respondents and trans men were insured (Table 1). Across the three gender identities surveyed, approximately 1 in 3 respondents indicated they had experienced difficulty accessing primary care (Figure 1A). For these respondents, their specific barriers were elicited (Figure 2). Among non-binary respondents (n=12), the most frequent barrier to primary care was fear of discrimination at the clinic.

**Table 1.**
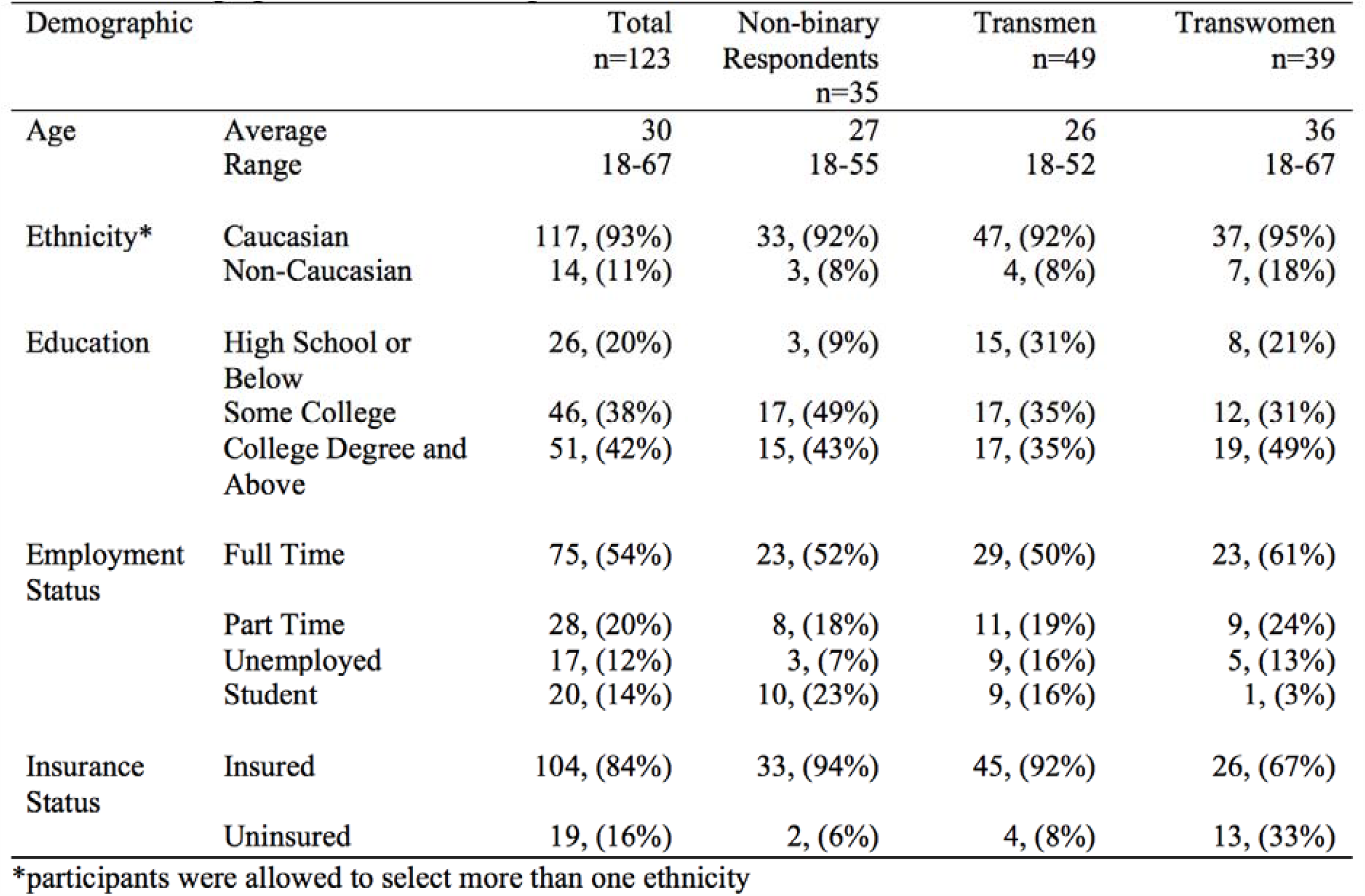
Demographic Data of the Respondents

| Demographic |  | Total<br>n=123 | Non-binary<br>Respondents<br>n=35 | Transmen<br>n=49 | Transwomen<br>n=39 |
| --- | --- | --- | --- | --- | --- |
| Age | Average | 30 | 27 | 26 | 36 |
|  | Range | 18-67 | 18-55 | 18-52 | 18-67 |
| Ethnicity* | Caucasian | 117, (93%) | 33, (92%) | 47, (92%) | 37, (95%) |
|  | Non-Caucasian | 14, (11%) | 3, (8%) | 4, (8%) | 7, (18%) |
| Education | High School or Below | 26, (20%) | 3, (9%) | 15, (31%) | 8, (21%) |
|  | Some College | 46, (38%) | 17, (49%) | 17, (35%) | 12, (31%) |
|  | College Degree and Above | 51, (42%) | 15, (43%) | 17, (35%) | 19, (49%) |
| Employment Status | Full Time | 75, (54%) | 23, (52%) | 29, (50%) | 23, (61%) |
|  | Part Time | 28, (20%) | 8, (18%) | 11, (19%) | 9, (24%) |
|  | Unemployed | 17, (12%) | 3, (7%) | 9, (16%) | 5, (13%) |
|  | Student | 20, (14%) | 10, (23%) | 9, (16%) | 1, (3%) |
| Insurance Status | Insured | 104, (84%) | 33, (94%) | 45, (92%) | 26, (67%) |
|  | Uninsured | 19, (16%) | 2, (6%) | 4, (8%) | 13, (33%) |

\*participants were allowed to select more than one ethnicity

**Figure 1.**
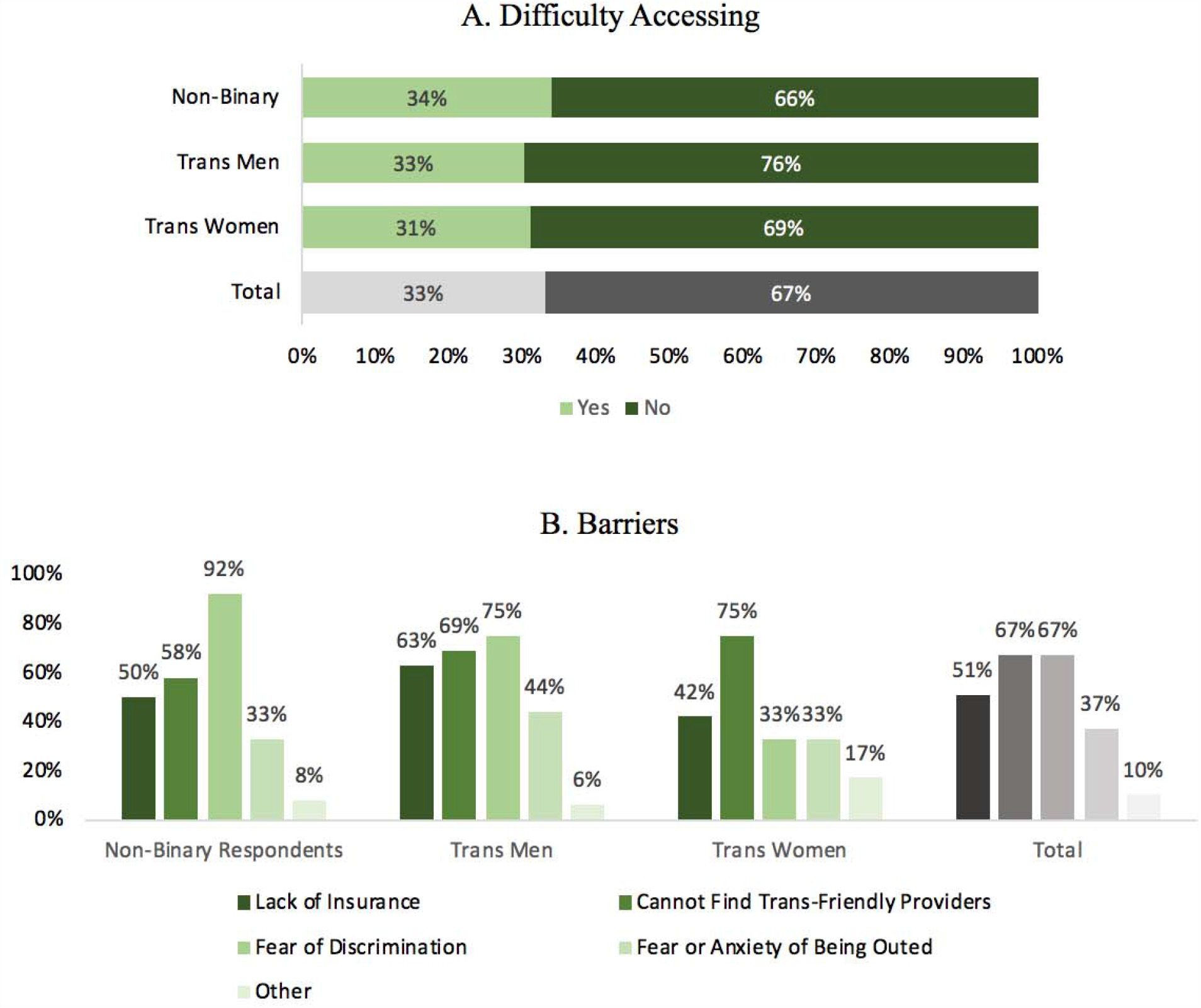
Primary Care. A. Difficulty Accessing. Each side bar represents a gender category: non-binary respondents (non-binary/ genderqueer/ agender/ transfeminine/ transmasculine), trans men (transgender female-to-male), trans women (transgender male-to-female), followed by the averaged totals of the 3 gender identities surveyed (in gray). B. Barriers. Each collection of bar graphs indicates a gender identity: non-binary respondents (non-binary/ genderqueer/ agender/ transfeminine/ transmasculine), trans men (transgender female-to-male), trans women (transgender male-to-female), followed by the averaged totals of the 3 gender identities surveyed (in gray). Each individual bar represents a barrier to primary care.

**Figure 2.**
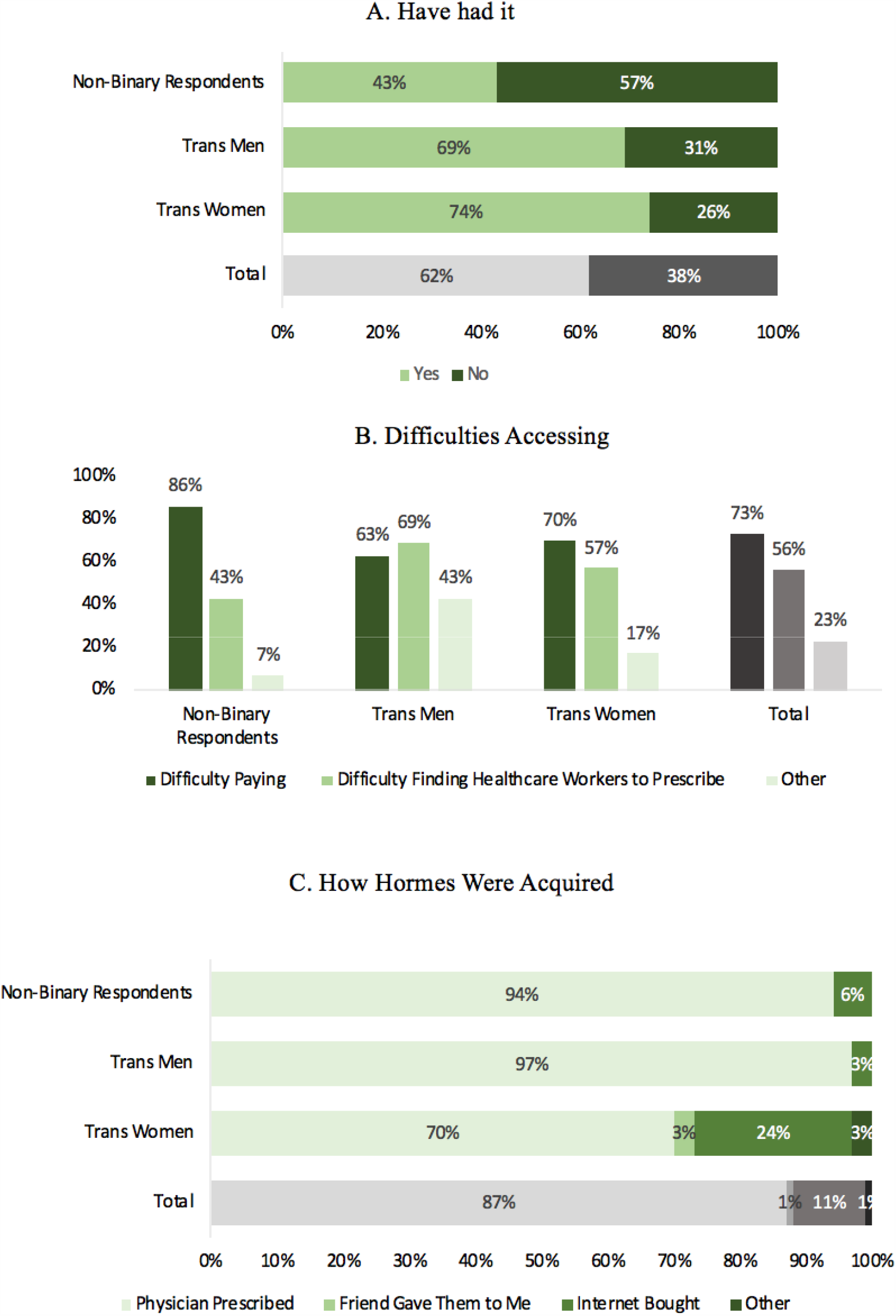
Hormone Therapy. A. Have had it. Each side bar represents a gender category: non-binary respondents (non-binary/ genderqueer/ agender/ transfeminine/ transmasculine), trans men (transgender female-to-male), trans women (transgender male-to-female), followed by the averaged totals of the 3 gender identities surveyed (in gray). B. Difficulties Accessing. Each collection of bar graphs indicates a gender identity: non-binary respondents (non-binary/ genderqueer/ agender/ transfeminine/ transmasculine), trans men (transgender female-to-male), trans women (transgender male-to-female), followed by the averaged totals of the 3 gender identities surveyed (in gray). Each individual bar represents a reason for difficulty in accessing hormones. C. How Hormones Were Acquired. Each side bar represents a gender category: non-binary respondents (non-binary/ genderqueer/ agender/ transfeminine/ transmasculine), trans men (transgender female-to-male), trans women (transgender male-to-female), followed by the averaged totals of the 3 gender identities surveyed (in gray). The color in the bar graph represents an option of where hormones were acquired.

Approximately half of the non-binary respondents indicated that insurance coverage and difficulty finding trans-friendly providers were major barriers to primary care. For trans men (n=16), the most frequently selected barrier was fear of discrimination (75%), followed closely by difficulty finding trans friendly providers (69%) and inadequate insurance coverage (63%). Forty-four percent of trans men selected fear of being outed (revealing trans status) as a barrier. Trans women (n=12) chose the difficulty finding trans-friendly providers as the most common barrier (75%), followed by the lack of insurance (45%) (Figure 1B).

The percent of respondents who have taken hormones varied by gender identity. Non-binary respondents indicated the lowest percentage of hormone therapy (43%). Both trans men and trans women reported a high frequency of hormone therapy (69% vs 74%, respectively). Overall, 62% were on hormone therapy (Figure 2A). Of the participants that reported receiving hormone therapy, additional data was collected regarding possible barriers to accessing it. Nearly 90% of the non-binary respondents (n=14) indicated they had difficulty paying for hormones, and nearly half indicated they had difficulty finding someone to prescribe hormone therapy. A majority of trans men (n=16) and trans women (n=23) also reported difficulty paying for hormones (63% and 70%) and finding a healthcare provider to prescribe this therapy (69% and 57%) (Figure 2B). Additional data was collected to determine how they accessed hormone therapy. Across each gender identity, the vast majority indicated hormone therapy was prescribed by a primary care physician (87%), but 24% of trans women indicated they acquired their hormones from the internet (Figure 2C).

Of the participants who reported not taking hormones, further data was collected regarding their interest in hormone therapy. Non-binary respondents (n=20) indicated the lowest percentage of interest (50%), whereas approximately 90% of trans men (n=15) and trans women (n=10) indicated interest in hormones (Figure 3A). Of the respondents who indicated they have not taken hormones, but responded they are interested in them, we queried their barriers to hormone therapy. One in two non-binary respondents (n=10) reported difficulty in finding providers with expertise in hormone therapy and cost as significant barriers. For trans men (n=14), 79% indicated cost as a barrier to hormones, and 64% listed lack of expert healthcare professionals as a barrier. Trans women (n=9) selected difficulty in finding a healthcare provider as a major barrier (84%), with other barriers selected at a much lower frequency. Overall, the most frequently selected barrier is the lack of professionals available or qualified to prescribe hormones (Figure 3C). Participants gave numerous comments regarding their experience accessing primary care (Table 2). These were sorted into three categories: difficulty accessing primary care, discrimination and lack of knowledge, and difficulty with cost and insurance.

**Table 3.**
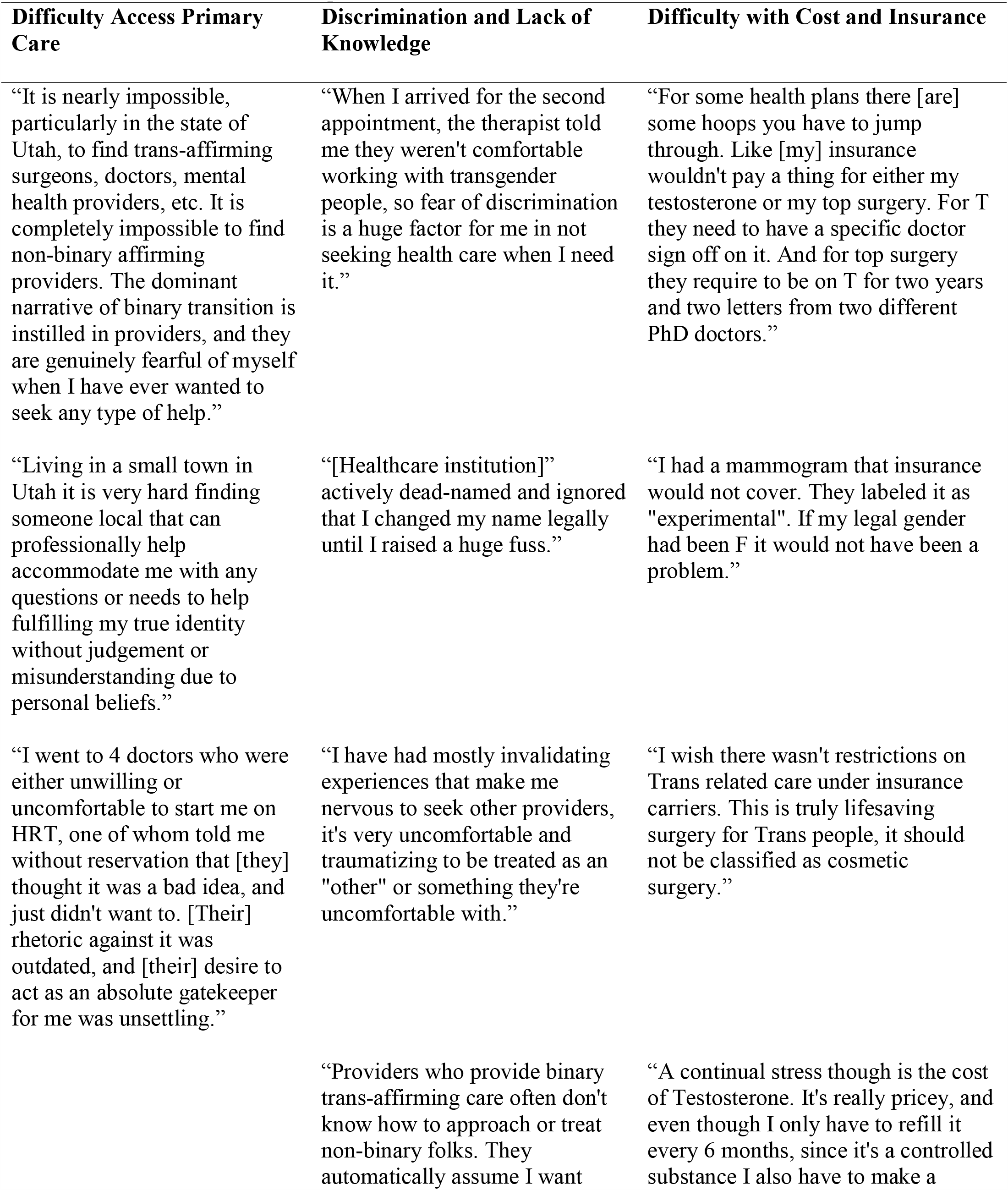
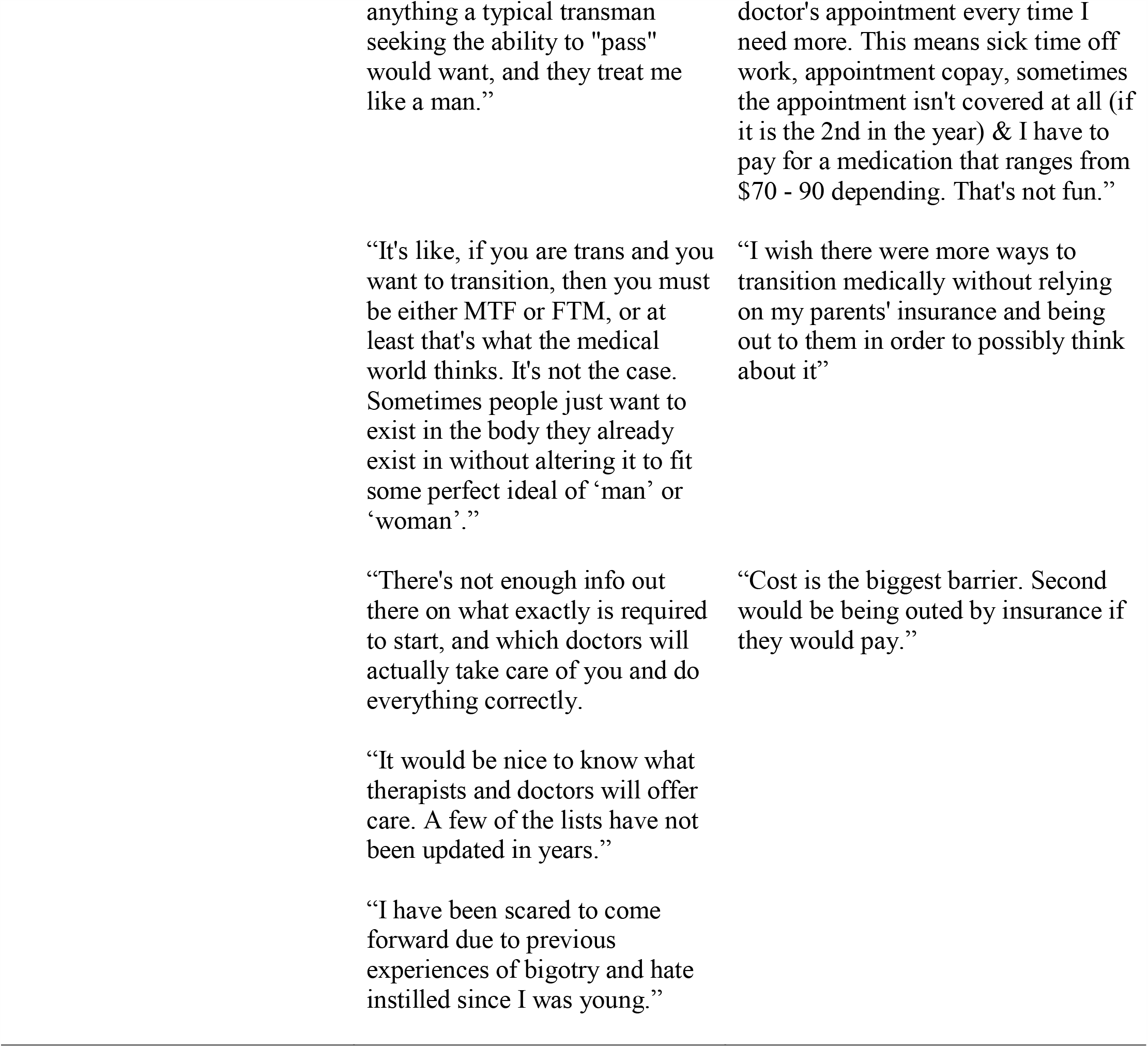
Comments from Respondents

**Figure 3.**
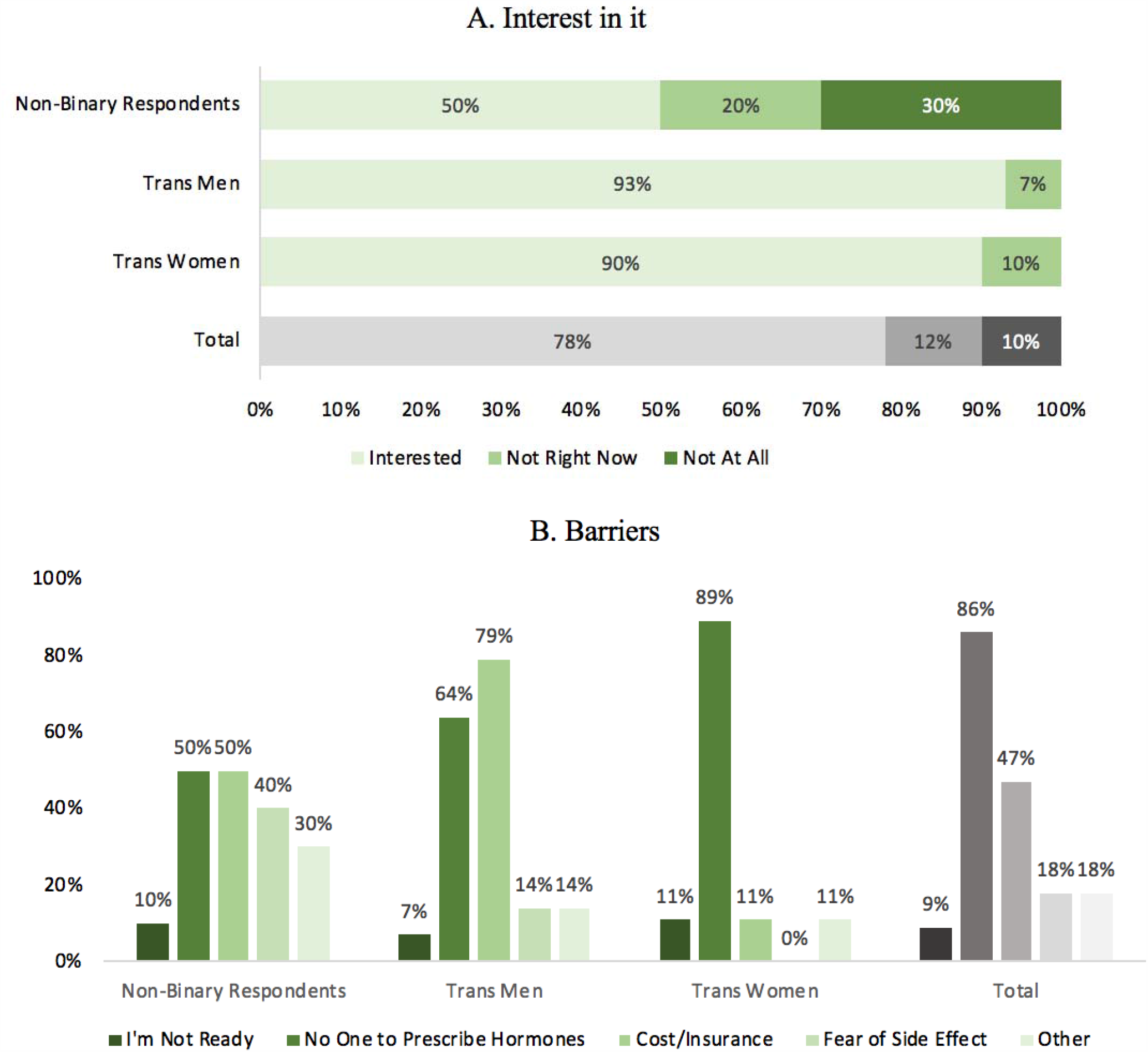
Respondents Who Have Not Had Hormone Therapy. A. Interest in it. Each side bar represents a gender category: non-binary respondents (non-binary/ genderqueer/ agender/ transfeminine/ transmasculine), trans men (transgender female-to-male), trans women (transgender male-to-female), followed by the averaged totals of the 3 gender identities surveyed (in gray). The color in the bar graph represents the level of interest in taking hormones, either interested, not interested right now, or not interested. B. Barriers. Each collection of bar graphs indicates a gender identity: non-binary respondents (non-binary/ genderqueer/ agender/ transfeminine/ transmasculine), trans men (transgender female-to-male), trans women (transgender male-to-female), followed by the averaged totals of the 3 gender identities surveyed (in gray). Each individual bar represents a barrier to accessing hormones.

## Discussion

### Difficulty Accessing Primary Care

Transgender and non-binary respondents in Utah indicated a 10-fold greater difficulty in finding a primary care physician compared to the general U.S. population: 30% as opposed to 3.2%.^12^ A contributing factor may well be that Utah is below the national average in density of registered nurses and has less than 1 physician per 1,000 people.^13,14^ Utah has a deficit of primary care providers relative to population, estimated to be a lack of 300 providers per 100,000.^18^ The lack of providers is further complicated by deficiency in primary care physicians that are trans-friendly and have the expertise to prescribe hormone therapy. The difficulty in finding primary care physicians who are trained for transgender healthcare is highlighted in the comments given by the participants. This issue, however, is not specific to Utah. Many transgender and non-binary people across the U.S have indicated that being unable to access trans-friendly providers is their largest barrier to care.^8^ For a liberal city such as New York, approximately 30% of transgender respondents reported having difficulty finding a trans-knowledgeable physician.^16^ Although their study was taken nearly 10 years ago, those numbers continue to reflect the percentage of transgender and non-binary residents of Utah who find that same difficulty. Over time, there has been an increase in transgender and non-binary medical training and awareness.^17^ However, our data suggests that Utah continues to lag behind in access and reflects what a liberal area looked like nearly a decade ago. The lack of adequately trained primary care providers in Utah comes with great consequences for transgender and non-binary individuals, not only for addressing basic healthcare needs, but also in accessing transition-specific healthcare. Nearly 90% of trans women who were not on hormone therapy listed being unable to find someone to prescribe hormones as a major barrier to accessing them. This lack of providers results in as many as 1 in 4 trans women who take hormones needing to find them online, and others getting them from friends, leading to potentially unsafe practices.

### Discrimination and Lack of Knowledge

The extremely high rate of fear of discrimination (67%) cited by respondents demonstrates this as a major barrier to accessing primary care and highlights a serious problem in the state of Utah that needs to be examined and corrected. Even if one has found a trans-friendly provider, patients could be anxious or worried about discrimination in the clinic or with the provider. Additionally, even if providers aren’t directly discriminatory, many healthcare workers aren’t trained to treat transgender and non-binary patients. This includes lack of knowledge regarding healthcare options or transition related care for those communities and being unaware of proper use of pronouns and preferred names. Many of the comments given by respondents (Table 2) reflect a fear of continued bigotry that prevents them from pursuing further healthcare. Previous literature suggests that for transgender and non-binary individuals surveyed in more liberal regions, the largest barrier to healthcare is access to a trans-friendly provider. Our data suggests that for a more conservative state such as Utah both the lack of trans-friendly providers and a fear of discrimination are large barriers. Nationally, 31% of trans men, 22% of trans women, and 20% of their non-binary respondents indicated fear of disrespect or mistreatment as a barrier to seeing a healthcare provider.^18^ Our data reflects that in Utah, non-binary individuals feel the highest percentage of fear of discrimination (32%), followed by trans men (25%) and trans women (11%) respectively. This suggests that discrimination in Utah is felt the strongest by non-binary individuals and trans men.

The data and comments overall continue to illustrate that it is not simply enough to express support for transgender and non-binary individuals; providers and their staff need to be educated and knowledgeable about the diverse experiences, desires, and identities of transgender and non-binary communities in Utah. Furthermore, many respondents expressed discontent at how few readily accessible resources are available for knowledge around transition or which providers offer transgender healthcare in Utah. The magnitude of negative and discriminatory healthcare experiences underscores the need for healthcare providers in Utah to improve outreach with LGBTQ+ organizations and advertise their services and support in more accessible ways.

### Difficulty with Cost and Insurance

The ability to pay for services is an added challenge for transgender and non-binary individuals, further complicated by the higher rates of unemployment faced by this community.^18^ While the majority of our respondents had some form of health insurance, it was apparent that for many, their policies did not pay for transition related care. For nearly half of the respondents who were interested in hormones but unable to get them, cost was listed as a major barrier. Many respondents detailed their experience and difficulty in paying for hormone therapy. Insurance costs come with an added stressor for transgender and non-binary people who are not out yet, given that respondents on their family’s insurance plans have to disclose their identity in order to access hormones through insurance. Given that approximately 1 in 3 participants listed “fear or anxiety of being outed” as a major barrier to accessing primary care, and many commented on their nervousness in speaking to their family about insurance for hormones, it is clear that in order to increase positive healthcare outcomes, the community in Utah must become more transgender-friendly. Moreover, there is evidence to suggest that covering transition-related healthcare decreases overall healthcare costs by reducing gender dysphoria, thereby reducing the need for mental health services and suicidal ideation.^19^ These observations highlight the urgent need to make primary care more accessible and reduce the discrimination felt by transgender and non-binary patients to achieve better health outcomes.

## Limitations

A major limitation is the small sample size which did not allow for statistically significant comparisons between the gender identities surveyed. The respondents predominantly identified as Caucasian, missing a large percentage of transgender and non-binary people of color. Survey-specific data collection has its own inherent limitations. There is no way to confirm the honesty of responses, the way in which questions were worded comes with slight bias, and multiple-choice options could miss a response or prompt a response that otherwise wouldn’t have been reported.

## Conclusion

Although Utah is a conservative state, its capitol city has a relatively high percentage of transgender and non-binary individuals. Results from this study confirm that transgender and non-binary adults in Utah have a 10 times harder time finding primary care physicians than the general population. One in three transgender and non-binary people have difficulty accessing the care they need. A combination of a lack of trans-friendly and trans-knowledgeable providers, fear of discrimination, difficulty with cost/insurance, and a lack of readily accessible knowledge makes it incredibly challenging for transgender and non-binary individuals to access healthcare in Utah. One in four trans women seek hormone therapy through the internet, leading to potentially unsafe medications. These observations suggest increased efforts to train community health care providers on trans-health and to work toward eliminating the culture of discrimination against the transgender and non-binary community. Furthermore, efforts are needed to advance anti-discriminatory laws within healthcare and insurance companies on the basis of gender identity and increase trust within the transgender and non-binary community so they can access the healthcare they need.

## Data Availability

The datasets generated and analyzed during the current study are not publicly available due to privacy of the participants.

## Abbreviations

None applicable

## Declarations

### Ethics approval and consent to participate

The University of Utah Institutional Review Board (review number 00105) declared that this study did not require formal ethics approval. Consent to participate in the study was obtained prior to participation.

### Consent for publication

Not applicable

### Authors’ contributions

BT analyzed and interpreted patient data. NM and JR were major contributors in writing of the manuscript. CA created and distributed the survey as well as aiding in interpretation of the data. All authors read and approved the final manuscript.

## Acknowledgements

Not applicable

